# Comparative effectiveness of ChAdOx1 versus BNT162b2 COVID-19 vaccines in Health and Social Care workers in England: a cohort study using OpenSAFELY

**DOI:** 10.1101/2021.10.13.21264937

**Authors:** William J Hulme, Elizabeth J Williamson, Amelia Green, Krishnan Bhaskaran, Helen I McDonald, Christopher T Rentsch, Anna Schultze, John Tazare, Helen J Curtis, Alex J Walker, Laurie Tomlinson, Tom Palmer, Elsie Horne, Brian MacKenna, Caroline E Morton, Amir Mehrkar, Louis Fisher, Seb Bacon, Dave Evans, Peter Inglesby, George Hickman, Simon Davy, Tom Ward, Richard Croker, Rosalind M Eggo, Angel YS Wong, Rohini Mathur, Kevin Wing, Harriet Forbes, Daniel Grint, Ian J Douglas, Stephen JW Evans, Liam Smeeth, Chris Bates, Jonathan Cockburn, John Parry, Frank Hester, Sam Harper, Jonathan AC Sterne, Miguel Hernán, Ben Goldacre

## Abstract

**Objectives:** To compare the effectiveness of the BNT162b2 mRNA (Pfizer-BioNTech) and the ChAdOx1 (Oxford-AstraZeneca) COVID-19 vaccines against infection and COVID-19 disease in health and social care workers.

**Design:** Cohort study, emulating a comparative effectiveness trial.

**Setting:** Linked primary care, hospital, and COVID-19 surveillance records available within the OpenSAFELY-TPP research platform.

**Participants:** 317,341 health and social care workers vaccinated between 4 January and 28 February 2021, registered with a GP practice using the TPP SystmOne clinical information system in England, and not clinically extremely vulnerable.

**Interventions:** Vaccination with either BNT162b2 or ChAdOx1 administered as part of the national COVID-19 vaccine roll-out.

**Main outcome measures:** Recorded SARS-CoV-2 positive test, or COVID-19 related Accident and Emergency attendance or hospital admission occurring within 20 weeks of vaccination.

**Results:** The cumulative incidence of each outcome was similar for both vaccines during the first 20 weeks post-vaccination. The cumulative incidence of recorded SARS-CoV-2 infection 6 weeks after vaccination with BNT162b2 was 19.2 per 1000 people (95%CI 18.6 to 19.7) and with ChAdOx1 was 18.9 (95%CI 17.6 to 20.3), representing a difference of -0.24 per 1000 people (95%CI -1.71 to 1.22). The difference in the cumulative incidence per 1000 people of COVID-19 accident and emergency attendance at 6 weeks was 0.01 per 1000 people (95%CI -0.27 to 0.28). For COVID-19 hospital admission, this difference was 0.03 per 1000 people (95%CI -0.22 to 0.27).

**Conclusions:** In this cohort of healthcare workers where we would not anticipate vaccine type to be related to health status, we found no substantial differences in the incidence of SARS-CoV-2 infection or COVID-19 disease up to 20 weeks after vaccination. Incidence dropped sharply after 3-4 weeks and there were very few COVID-19 hospital attendance and admission events after this period. This is in line with expected onset of vaccine-induced immunity, and suggests strong protection against COVID-19 disease for both vaccines.

## Introduction

The COVID-19 global pandemic has prompted the rapid development and delivery of vaccines to combat the disease. Following demonstration of high safety and efficacy against symptomatic and severe disease in phase-III randomised controlled trials (RCTs), two vaccines have been approved and widely administered as part of the national vaccination programme in the United Kingdom: the Pfizer-BioNTech BNT162b2 mRNA COVID-19 vaccine (1) (*BNT162b2*) and the Oxford-AstraZeneca ChAdOx1 nCoV-19 vaccine (2) (*ChAdOx1*). Post-authorisation assessment of vaccine effectiveness using observational data is necessary to monitor the success of such programmes as, invariably, target populations and settings differ substantially from those of trials. To date, there have been no RCTs that have directly compared the BNT162b2 and ChAdOx1 vaccines to estimate the relative efficacy against COVID-19 infection and disease in the same population. The concurrent roll-out of these vaccines across the UK (3), combined with the country’s well-developed infrastructure for conducting research using routinely-collected primary care health data, provides a rare opportunity to emulate such a trial using observational data.

COVID-19 vaccination in the UK has been prioritised based on the risk of infection and subsequent severity of disease (4). Health and social care workers (HCWs) were amongst the first groups eligible for vaccination due to the high occupational exposure to the SARS-CoV-2 virus, making HCW status an important confounder of the effect of vaccination on infection and post-infection disease. By studying this group in isolation, this confounding is mitigated, and vaccine effects in a healthy, high-exposure group can be estimated.

This cohort study used a target-trial design to assess the effectiveness of ChAdOx1 compared with BNT162b2 in HCWs, including second dose effectiveness, using the OpenSAFELY-TPP linked primary care database covering around 40% of England’s population.

## Methods

### Data source

The OpenSAFELY-TPP database (https://opensafely.org) covers 24 million people registered at GP practices that use TPP SystmOne electronic health record software. It includes pseudonymized data such as coded diagnoses, medications and physiological parameters. No free text data are included. This primary care data is linked (via NHS numbers) with A&E attendance and in-patient hospital spell records via NHS Digital’s Hospital Episode Statistics (HES), national coronavirus testing records via the Second Generation Surveillance System (SGSS), and national death registry records from the Office for National Statistics (ONS). Vaccination status is available in the GP record directly via the National Immunisation Management System (NIMS). HCW status is recorded for all vaccine recipients at the time of vaccination, and this information is sent to OpenSAFELY-TPP from NHS Digital’s COVID-19 data store.

### Study population

We studied health and social care workers in England vaccinated with either BNT162b2 or ChAdOx1. This group was prioritised for vaccination at the start of the vaccine roll-out due to the high occupational exposure to the SARS-CoV-2 virus, and many were vaccinated during the period where both vaccines were widely used.

Vaccinated HCWs were included in the study if: they were registered at a GP practice using TPP’s SystmOne clinical information system on the day that they received their first dose of BNT162b2 or ChAdOx1; the date of vaccination was between 04 January (when ChAdOx1 was first administered in England) and 28 February 2021, a 56-day period during which both vaccines were being administered widely; they were aged between 18 and 64 inclusive; not classed as Clinically Extremely Vulnerable, as set out by government guidance (4), at the time of vaccination; information on sex, ethnicity, deprivation, and geographical region was known. Study participants were followed up for no more than 20 weeks from the day of the first dose, including time after their second dose. Follow-up was censored earlier than this at 13 June 2021, death, or de-registration.

### Outcomes

Three outcomes were defined: positive SARS-CoV-2 test; COVID-19 A&E attendance; and unplanned COVID-19 hospital admission. Positive SARS-CoV-2 tests were identified using SGSS records and based on swab date. Both polymerase chain reaction (PCR) and lateral flow tests are included, without differentiation between symptomatic and asymptomatic infection. COVID-19 A&E attendances were identified using HES emergency care records with U07.1 (“COVID-19, virus identified”) or U07.2 (“COVID-19, virus not identified”) ICD-10 diagnosis codes (5). Unplanned COVID-19 hospital admissions were identified using HES in-patient hospital records with U07.1 or U07.2 reason for admission ICD-10 codes.

Although severe disease (such as requirement for intensive or critical care) and mortality were of interest there were too few events to investigate these outcomes fully. Unadjusted incidence of COVID-19 deaths are reported descriptively. These were identified using linked death registration data. Deaths COVID-19 ICD-10 codes (as above) mentioned anywhere on the death certificate (i.e., as an underlying or contributing cause of death) were included.

### Additional variables

Participant characteristics used describe the cohort and for confounder adjustment include: age, sex (male or female), English Index of Multiple Deprivation (IMD, grouped by quintiles), ethnicity (Black, Mixed, South Asian, White, Other, as per the UK census), NHS region (East of England, Midlands, London, North East and Yorkshire, North West, South East, South West), number of conditions in the clinically “at risk” (but not clinically extremely vulnerable) classification, as per national prioritisation guidelines, the number of SARS-CoV-2 tests in the 90 days prior to the study start date (via SGSS), rurality (urban conurbation, urban city or town, rural town or village), evidence of prior SARS-CoV-2 infection (positive test or COVID-19 hospitalisation), learning disabilities, severe mental illness. All characteristics were ascertained as at the time of vaccination.

### Statistical Analysis

We compared the effectiveness of a first dose of ChAdOx1 versus BNT162b2 using pooled logistic regression (6) (7) (PLR) on the time since vaccination timescale with the outcome risk estimated each day. The effect is permitted to vary over the timescale to account for the potential time-varying differences in vaccine protection between the two brands. A PLR model can be used to approximate Cox models with time-varying treatment effects on the time-since-vaccination timescale, and enables the estimation of risk-adjusted cumulative incidence for each vaccine type. This is the average over all participants of the cumulative incidence for each day of follow-up predicted by the PLR model, under the (counterfactual) assumption that everyone received the BNT162b2 vaccine or that everyone received the ChAdOx1 vaccine. Assuming adequate confounder adjustment, this estimates the vaccine-specific cumulative incidence that would have been observed in an RCT comparing the two vaccines in the population under consideration. Standard errors for the PLR model were obtained using the clustered sandwich estimator to account for within-participant clustering. Confidence intervals for the vaccine-specific marginal cumulative incidence, and their difference, were obtained using the delta method (i.e, a first-order Taylor series approximation of the variance).

The PLR model included vaccine type, and a vaccine-specific 3-knot restricted cubic spline for time since vaccination. Knot locations were based on quartiles of the event times. Three models were fit for each outcome, with progressive adjustment for confounders: (1) adjusting for region-specific calendar-time effects, by including a 2-knot restricted cubic spline for the date of vaccination and its interaction with region; (2) additionally adjusting for demographic characteristics; (3) additionally adjusting for clinical characteristics. During the study period, the advice from the UK Chief Medical Officers was that “second doses of both vaccines will be administered towards the end of the recommended vaccine dosing schedule of 12 weeks” (8). Using an intention-to-treat approach, comparative effectiveness estimates beyond 14 weeks were considered to be second-dose effects. We report the actual timing of second doses to assess the extent of any deviation from this recommended treatment strategy, frequency of any cross-brand second doses, and how these may differ between vaccine types.

PLR models are computationally expensive to fit, as the input dataset must be arranged as one row per person per day of follow-up. To manage this, a sampling strategy was used such that all those who experienced the event of interest are selected (to retain statistical power), and a random sample of 50,000 event-free participants are selected. Person-time is weighted by the inverse of the sampling probability to recover the characteristics of the complete cohort.

### Missing data

A complete-case approach was used to deal with missing values. After exclusions for missing values on demographic variables (see exclusion criteria), there were no missing values in remaining variables as they were defined by the presence or absence of clinical codes or events.

### Software, code, and reproducibility

Data management and analyses were conducted in Python 3.8 and R version 4.0.2. All code is available at https://github.com/opensafely/comparative-ve-research, and is shared openly for review and re-use under MIT open license. Codelists are available at https://www.opencodelists.org/. No person-level data is shared. Any reported figures based on counts below 6 are redacted or rounded for disclosure control. This study followed the STROBE-RECORD reporting guidelines.

### Patient and public involvement

We have developed a publicly available website https://opensafely.org/ through which we invite any patient or member of the public to contact us regarding this study or the broader OpenSAFELY project.

## Results

### Study population

A total of 361,287 HCWs aged 18-64 receiving a first dose of BNT162b2 or ChAdOx1 between 4 January and 28 February 2021 and actively registered at at TPP practice were identified, with 317,341 (87.8%) meeting the study eligibility criteria (supplementary Tables S1 and S2).

In total, 253,134 (79.8%) were vaccinated with BNT162b2, contributing 95,420 person-years of potential follow-up (including all person-time prior to a censoring event, and possibly after an outcome event). For ChAdOx1, this was 64,207 (20.2%) and 23,351 person-years.

Characteristics were largely well-balanced between recipients of each vaccine, though regional and temporal differences in the distribution of each vaccine are notable (Table 1). BNT162b2 was on average administered earlier than ChAdOx1 (Figure 1; median day of vaccination 15 January for BNT162b2, 22 January for ChadOx1). BNT162b2 was relatively more likely to be administered in the South and East of England, and ChAdOx1 the Midlands and Northern England. Evidence of prior SARS-CoV-2 infection was higher in ChAdOx1 recipients (10.8% for BNT162b2, 14.1% for ChAdOx1), consistent with ChAdOx1 recipients being vaccinated later on average. The proportion of each clinical condition is slightly higher in ChAdOx1 recipients, though consistently under a 0.6% percent-point difference.

**Table 1:** Baseline characteristics. Participant characteristics as on the day of vaccination.

| Characteristic | BNT162b2 | ChAdOx1 |
| --- | --- | --- |
| Total N | 253,134 | 64,207 |
| Age |  |  |
| 18-30 | 41,086 (16%) | 10,464 (16%) |
| 30s | 59,518 (24%) | 14,724 (23%) |
| 40s | 64,553 (26%) | 16,286 (25%) |
| 50s | 67,776 (27%) | 17,543 (27%) |
| 60-64 | 20,201 (8.0%) | 5,190 (8.1%) |
| Sex |  |  |
| Female | 200,149 (79%) | 49,331 (77%) |
| Male | 52,985 (21%) | 14,876 (23%) |
| Ethnicity |  |  |
| White | 211,463 (84%) | 54,287 (85%) |
| Black | 8,518 (3.4%) | 3,161 (4.9%) |
| South Asian | 23,140 (9.1%) | 4,701 (7.3%) |
| Mixed | 3,848 (1.5%) | 1,007 (1.6%) |
| Other | 6,165 (2.4%) | 1,051 (1.6%) |
| IMD |  |  |
| 1 most deprived | 36,850 (15%) | 10,300 (16%) |
| 2 | 47,279 (19%) | 12,116 (19%) |
| 3 | 55,832 (22%) | 13,751 (21%) |
| 4 | 57,499 (23%) | 14,353 (22%) |
| 5 least deprived | 55,674 (22%) | 13,687 (21%) |
| Region |  |  |
| North East and Yorkshire | 53,522 (21%) | 16,418 (26%) |
| East of England | 62,377 (25%) | 11,861 (18%) |
| Midlands | 50,582 (20%) | 17,224 (27%) |
| South West | 35,948 (14%) | 5,056 (7.9%) |
| London | 10,405 (4.1%) | 2,148 (3.3%) |
| North West | 23,644 (9.3%) | 7,875 (12%) |
| South East | 16,656 (6.6%) | 3,625 (5.6%) |
| Rural/urban category |  |  |
| Urban conurbation | 61,699 (24%) | 19,295 (30%) |
| Urban city or town | 144,222 (57%) | 32,194 (50%) |
| Rural town or village | 47,213 (19%) | 12,718 (20%) |
| Vaccination day (from 4 January 2021) | 12 (7, 18) | 19 (13, 33) |
| Body Mass Index > 40 kg/m<sup>2</sup> | 9,789 (3.9%) | 2,860 (4.5%) |
| Chronic heart disease | 9,207 (3.6%) | 2,527 (3.9%) |
| Chronic kidney disease | 1,994 (0.8%) | 551 (0.9%) |
| Diabetes | 12,674 (5.0%) | 3,339 (5.2%) |
| Chronic liver disease | 3,854 (1.5%) | 1,189 (1.9%) |
| Chronic respiratory disease | 2,579 (1.0%) | 731 (1.1%) |
| Chronic neurological disease | 6,063 (2.4%) | 1,620 (2.5%) |
| Immunosuppressed | 2,527 (1.0%) | 681 (1.1%) |
| Asplenia or poor spleen function | 1,704 (0.7%) | 481 (0.7%) |
| Learning disabilities | 187 (<0.1%) | 60 (<0.1%) |
| Serious mental illness | 1,276 (0.5%) | 434 (0.7%) |
| Morbidity count |  |  |
| 0 | 210,107 (83%) | 52,455 (82%) |
| 1 | 36,544 (14%) | 9,835 (15%) |
| 2+ | 6,483 (2.6%) | 1,917 (3.0%) |
| Prior SARS-CoV-2 infection | 27,312 (11%) | 9,085 (14%) |
| Number of SARS-CoV-2 tests in previous 3 months |  |  |
| 0 | 160,576 (63%) | 42,095 (66%) |
| 1-3 | 73,611 (29%) | 18,946 (30%) |
| 4-6 | 10,696 (4.2%) | 1,765 (2.7%) |
| 7+ | 8,251 (3.3%) | 1,401 (2.2%) |

**Figure 1:**
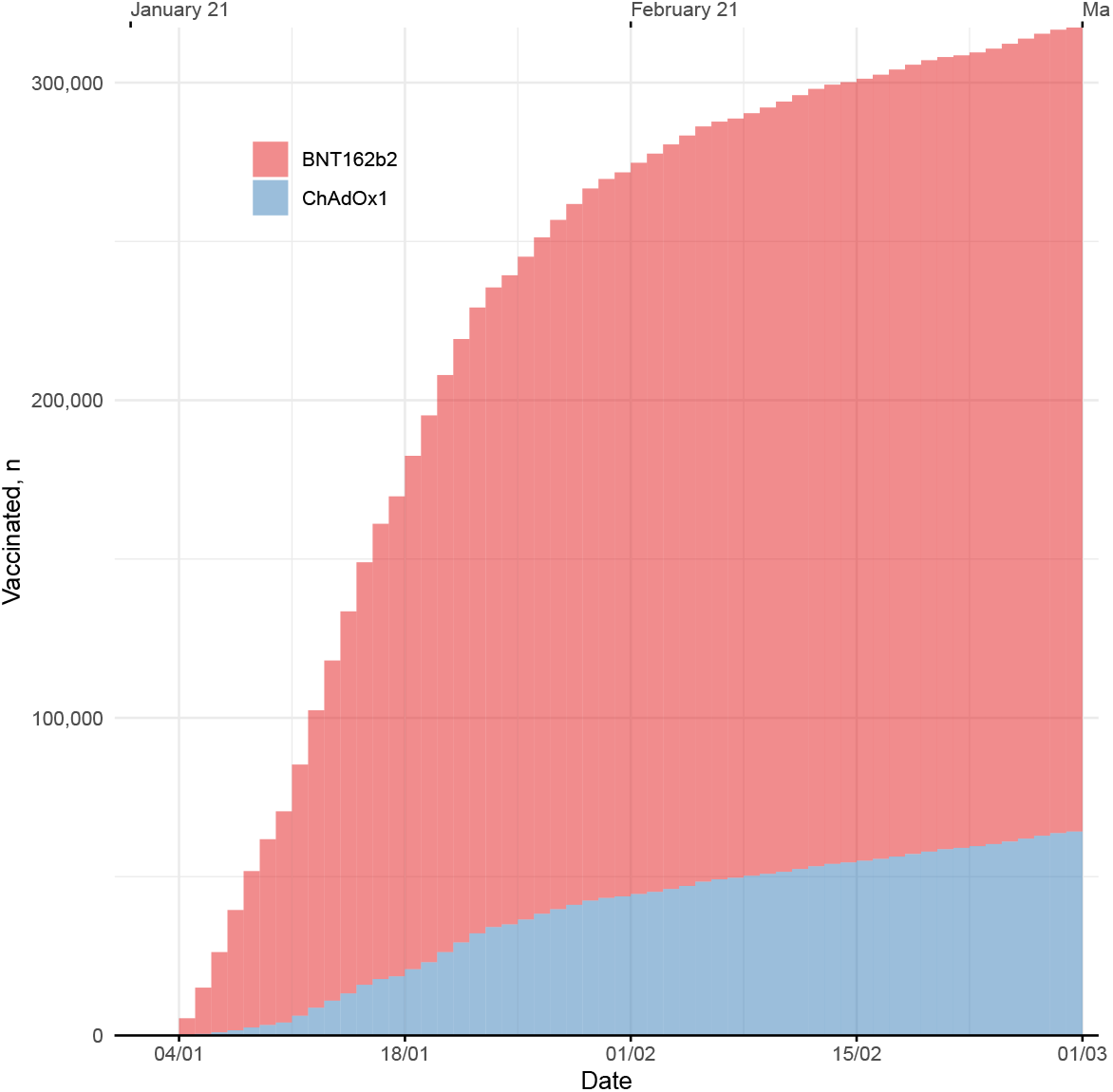
Vaccination dates. Cumulative enrollment of study participants by date.

### Events

Over the duration of 118,771 person-years of follow-up there were 6,962 positive SARS-CoV-2 tests, 282 COVID-19 A&E attendances, and 166 COVID-19 hospital admissions. While not a primary outcome, there were also 47 deaths from any cause, of which fewer than six were COVID-19 related (supplementary Table S3).

At 12 weeks, 95.4% of BNT162b2 recipients had received a second dose, compared with 90.8% of ChAdOx1 recipients. There were 413 (0.13%) participants who received a second dose within 20 weeks that was not the same brand as the first dose, including Moderna mRNA-1273 vaccine second doses (supplementary Figure S3, Table S4).

### Comparative effectiveness

At 6 weeks post-vaccination, the ChAdOx1 versus BNT162b2 absolute risk difference per 1000 people (Figure 2) for a positive SARS-CoV-2 test was -0.24 (95%CI -1.71 to 1.22), for COVID-19 A&E attendances was 0.01 (−0.27 to 0.28) and for COVID-19 hospital admissions was 0.03 (−0.22 to 0.27).

**Figure 2:**
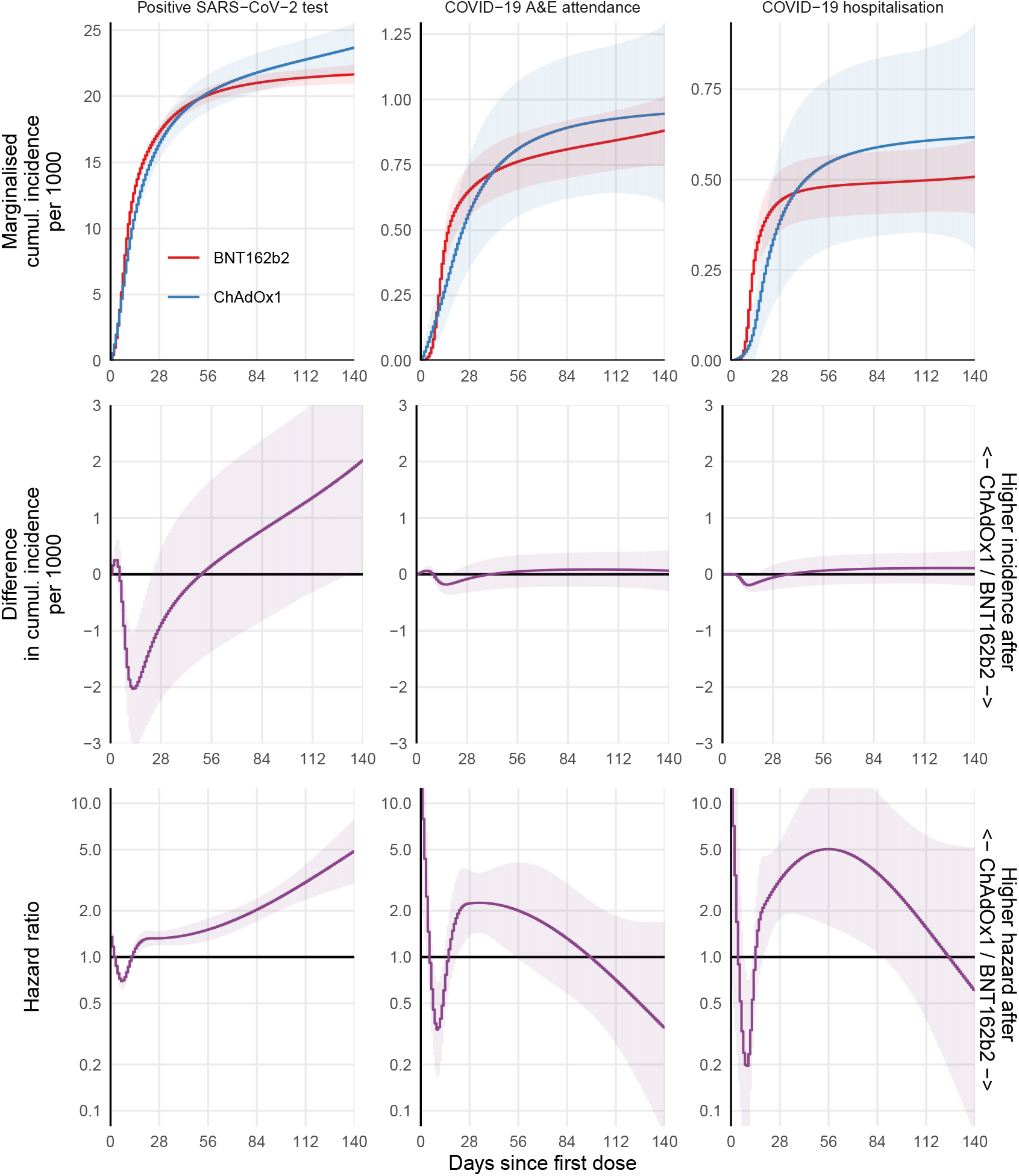
Comparative effectiveness. For each outcome based on the fully adjusted model, the marginal cumulative incidence for ChAdOx1 and BNT162b2, their difference, and the hazard ratio are shown. Models that assumed piecewise-constant hazards gave similar effect estimates (supplementary Figure S2). The models with less extensive confounder adjustment gave very similar estimates (supplementary Figure S1) suggesting that recipients of each vaccine were similar after accounting for differences in vaccine allocation over space and time (as did all models).

At 18 weeks post-vaccination, the absolute risk difference per 1000 people for a positive SARS-CoV-2 test was 1.69 (95%CI -0.09 to 3.46), for COVID-19 A&E attendances was 0.08 (−0.26 to 0.41) and for COVID-19 hospital admissions was 0.11 (−0.18 to 0.40).

The estimated log hazard ratios were non-zero at certain periods (Figure 2), but the underlying absolute risk was small enough that this did not manifest in substantial differences in the cumulative incidence beyond the fourth week.

## Discussion

This observational study of 317,341 health and social care workers living in England found similar outcomes for those receiving the BNT162b2 or ChAdOx1 COVID-19 vaccines during the first 20 weeks post-vaccination. There is a clear leveling-off of the incidence of positive tests after around 3-4 weeks for both vaccines, consistent with the expected time-to-onset of vaccine-induced immunity of around 2 weeks plus the delay from infection to a positive test. At 20 weeks, the difference in the cumulative incidence of positive test was 2.04 per 1000 people (0.04, 4.04) higher for ChAdOx1 compared with BNT162b2, and extrapolating beyond this time would suggest a small advantage for BNT162b2. However, BNT162b2 recipients were more likely to receive their second dose sooner, conferring a greater protective effect that is not accounted for in this estimate.

There were very few COVID-19 hospital attendance and admission events after this period, and the difference in the cumulative incidence (and their CIs) was well below 1 event per 1000 people in either direction.

### Strengths and weaknesses

We used routinely-collected health records with comprehensive coverage of primary care, hospital admissions, COVID-19 testing, COVID-19 vaccination, and death registrations to study vaccinated health and social care workers. This group were eligible for vaccination at the start of the UK’s vaccination programme due to exposure to higher viral loads and the need to reduce enforced absences in essential healthcare workers during a global pandemic. As such, HCW status is an important determinant of both vaccination time and SARS-CoV-2 infection, and a certain source of confounding in studies of vaccine effectiveness which may be reduced by studying this group in isolation. Although it is not typically possible to identify HCWs in NHS primary care records, this information was comprehensively collected at the time of COVID-19 vaccination and so is known for all vaccine recipients. HCWs are the only early vaccinees who are relatively young and healthy, and were vaccinated during a period where infection rates were high and both vaccines were being widely administered (3). Overall, this provides a rare opportunity to study comparative effectiveness under conditions that, to some extent, approximate random vaccine allocation. However, some limitations remain.

Despite reasonable balance of vaccine allocation across baseline characteristics and adjustment for a range of potential confounders, the possibility of unmeasured confounding remains. The cold storage requirements of BNT162b2 meant that it was more likely to have be administered in Acute NHS Trusts and other large vaccination centres, and is thus a potential confounder for instance due to differences in viral exposure across these settings. Though we adjusted for region, rurality, and deprivation, we were unable to directly account for occupational differences that may affect both exposure risk and vaccine type. We note also that the GP practices covered in this study are not geographically representative, as TPP SystmOne software is not widely-used in some regions such as London and the North West.

We were unable to fully investigate differences in protection against severe disease, in large part due to clear protective benefits of both vaccines, reducing the absolute numbers of events, and therefore statistical power, in the studied cohort.

Second-dose effects were considered using an intention-to-treat approach, which does not account for potential differences in the timing of the second dose between vaccine types. However, we found that by 12 weeks more BNT162b2 recipients had received a second dose than ChAdOx1 recipients, which would bias effect estimates in favour of BNT162b2 assuming higher protection after a second-dose of either vaccine. Regardless, there was insufficient power to reliably estimate comparative second-dose effects as event incidence at 12 weeks and beyond was low for both vaccines, with fewer than 10 positive tests per 1000-person-years, and far fewer for COVID-19 A&E attendances and hospital admissions during this period. The need for many thousands more person-years to detect differences in second-dose effects is encouraging to the extent that it suggests extremely high protective effects for both vaccines, though the fact that background case rates in England were declining over the duration of the study period should not be ignored.

### Findings in context

A number of studies have estimated COVID-19 vaccine effectiveness in observational data (9) (10) (11) (12). Vaccine protection is typically assessed by comparing vaccinated against unvaccinated person-time, or against the early post-vaccination period before anticipated onset of vaccine-induced immunity. Such designs are vulnerable to confounding and selection bias, for example due to vaccine prioritisation and eligibility policies. In addition, differences in vaccine access and acceptance can cause substantial imbalance between vaccinated and unvaccinated groups that may be impossible to account for. Many of these biases can be bypassed when making direct comparisons between recipients of different vaccine types, if the relative performance of two vaccines is of interest. To our knowledge, the present study is the first to assess effectiveness of the BNT162b2 and ChAdOx1 vaccines in a head-to-head comparison in either an experimental or observational setting.

Our study suggests that there is little to differentiate the vaccines with respect to their respective protective effects against infection and hospital-related COVID-19 outcomes. This pushes decisions about which vaccine to favour onto other attributes such as safety profile, logistics, and cost. However, we cannot rule-out larger differences in effectiveness in other settings. For instance: results seen in this cohort of predominantly healthy HCWs may not reflect comparative effectiveness in more vulnerable groups, such as the elderly or immunosuppressed; the dominant circulating variant during the period of study was the Alpha variant but has now been replaced by Delta variant against which vaccines may be less effective (13); longer-term effectiveness, and in particular the potential immunological waning, may differ.

## Conclusion

This study found no substantial differences in the incidence of SARS-CoV-2 infection or COVID-19-related hospital events following vaccination with BNT162b2 or ChAdOx1 in a cohort of health and social care workers in England. The incidence of COVID-19-related A&E attendances or hospital admissions four weeks after the first dose of either vaccine was fewer than one event in 10,000-person-years, suggesting strong protective effects for both vaccines but nevertheless limiting power to reliably estimate comparative effectiveness with respect to severe COVID-19 outcomes. Further studies are needed to assess comparative effectiveness in newer, more prevalent variants and to assess longer-term effectiveness.

## Summary

### What is already known

- Phase III randomised controlled trials show clear evidence of efficacy of the BNT162b2 mRNA and the ChAdOx1 vaccines
- Observational studies have provided further evidence of effectiveness in mass vaccine rollout settings.
- No head-to-head comparison of these vaccines has been made in randomised trials, and such comparisons are difficult in observational data without administration of both vaccines across the same population at the same time.

### What this study adds

- By exploiting the concurrent rollout of both vaccines in health and social care workers in England, we were able to make direct comparisons of first and second dose effectiveness between the vaccines.
- We found similar incidence rates for infection and COVID-19-related hospital attendance and admission.

## Data Availability

All code is available at https://github.com/opensafely/comparative-ve-research, and is shared openly for review and re-use under MIT open license. Codelists are available at https://www.opencodelists.org/. No person-level data is shared. Any reported figures based on counts below 6 are redacted or rounded for disclosure control.

## Acknowledgements

We are very grateful for all the support received from the TPP Technical Operations team throughout this work, and for generous assistance from the information governance and database teams at NHS England / NHSX.

## Funding

This work was supported by the Medical Research Council MR/V015737/1 and the Longitudinal Health and wellbeing strand of the National Core Studies programme. The OpenSAFELY platform is funded by the Wellcome Trust. TPP provided technical expertise and infrastructure within their data centre pro bono in the context of a national emergency. BGs work on clinical informatics is supported by the NIHR Oxford Biomedical Research Centre and the NIHR Applied Research Collaboration Oxford and Thames Valley. BG’s work on better use of data in healthcare more broadly is currently funded in part by: NIHR Oxford Biomedical Research Centre, NIHR Applied Research Collaboration Oxford and Thames Valley, the Mohn-Westlake Foundation, NHS England, and the Health Foundation; all DataLab staff are supported by BG’s grants on this work. LS reports grants from Wellcome, MRC, NIHR, UKRI, British Council, GSK, British Heart Foundation, and Diabetes UK outside this work. KB holds a Wellcome Senior Research Fellowship (220283/Z/20/Z). HIM is funded by the NIHR Health Protection Research Unit in Immunisation, a partnership between Public Health England and London School of Hygiene & Tropical Medicine. AYSW holds a fellowship from the British Heart Foundation. EJW holds grants from MRC. RM holds a Sir Henry Wellcome Fellowship funded by the Wellcome Trust (201375/Z/16/Z). HF holds a UKRI fellowship. IJD holds grants from NIHR and GSK.

Funders had no role in the study design, collection, analysis, and interpretation of data; in the writing of the report and in the decision to submit the article for publication.

The views expressed are those of the authors and not necessarily those of the NIHR, NHS England, Public Health England or the Department of Health and Social Care.

For the purpose of Open Access, the author has applied a CC BY public copyright licence to any Author Accepted Manuscript (AAM) version arising from this submission.

### Information governance and ethical approval

NHS England is the data controller; TPP is the data processor; and the key researchers on OpenSAFELY are acting with the approval of NHS England. This implementation of OpenSAFELY is hosted within the TPP environment which is accredited to the ISO 27001 information security standard and is NHS IG Toolkit compliant (14) (15) ; Patient data has been pseudonymised for analysis and linkage using industry standard cryptographic hashing techniques; all pseudonymised datasets transmitted for linkage onto OpenSAFELY are encrypted; access to the platform is via a virtual private network (VPN) connection, restricted to a small group of researchers; the researchers hold contracts with NHS England and only access the platform to initiate database queries and statistical models; all database activity is logged; only aggregate statistical outputs leave the platform environment following best practice for anonymisation of results such as statistical disclosure control for low cell counts (16). The OpenSAFELY research platform adheres to the obligations of the UK General Data Protection Regulation (GDPR) and the Data Protection Act 2018. In March 2020, the Secretary of State for Health and Social Care used powers under the UK Health Service (Control of Patient Information) Regulations 2002 (COPI) to require organisations to process confidential patient information for the purposes of protecting public health, providing healthcare services to the public and monitoring and managing the COVID-19 outbreak and incidents of exposure; this sets aside the requirement for patient consent (17). Taken together, these provide the legal bases to link patient datasets on the OpenSAFELY platform. GP practices, from which the primary care data are obtained, are required to share relevant health information to support the public health response to the pandemic, and have been informed of the OpenSAFELY analytics platform.

This study was approved by the Health Research Authority (REC reference 20/LO/0651) and by the LSHTM Ethics Board (reference 21863).

### Guarantor

BG is guarantor.

