## Supplement for "Comparative effectiveness of ChAdOx1 versus BNT162b2 COVID-19 vaccines in Health and Social Care workers in England: a cohort study using OpenSAFELY"

### Comparative vaccine effectiveness of ChAdOx1 versus BNT162b2 in Health and Social Care workers in England: Supplementary materials

**Figure S1a: Comparative effectiveness adjusting for region-specific calendar-time**

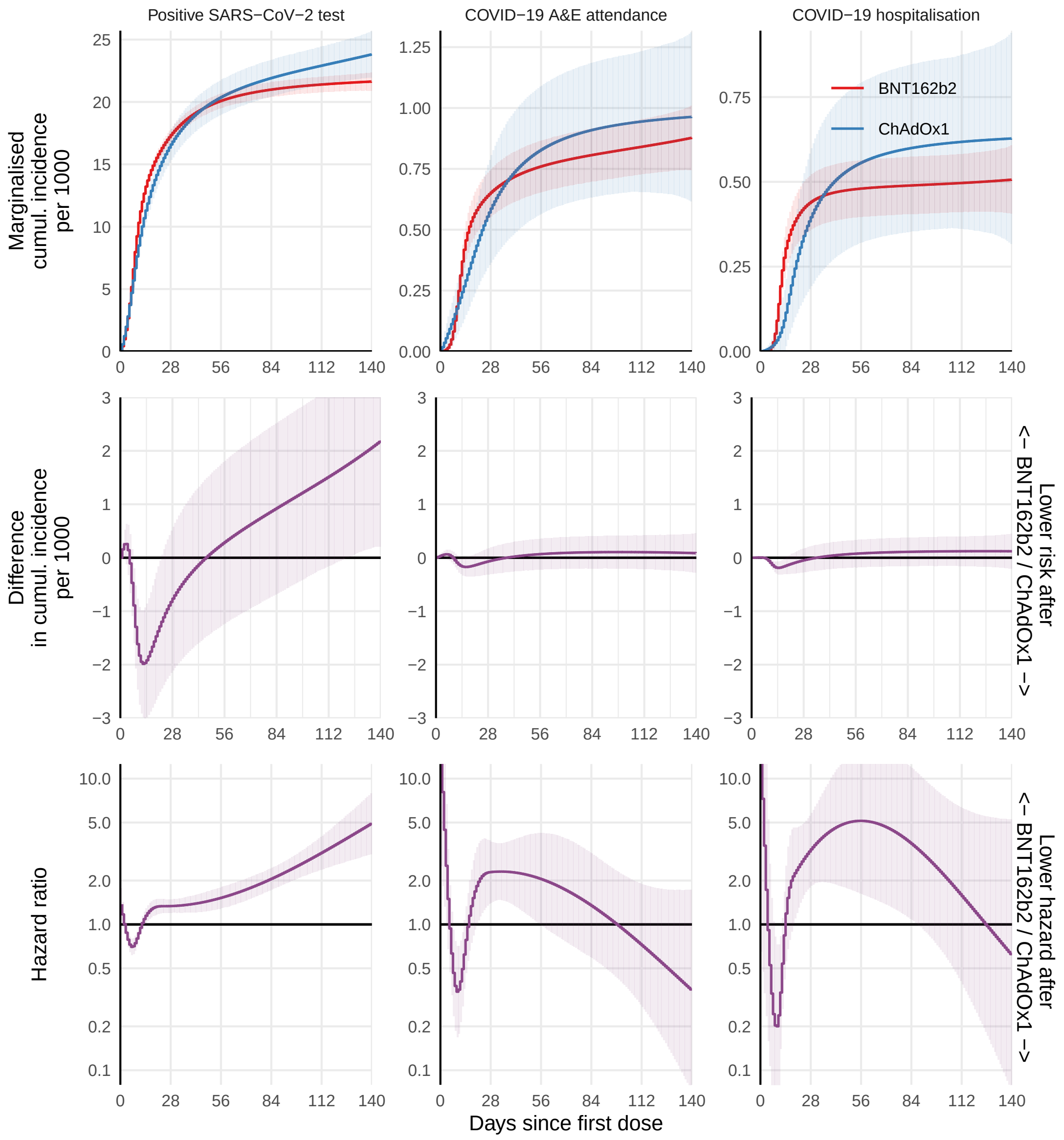

**Figure S1b: Comparative effectiveness adjusting for region-specific calendar-time and demographic characteristics**

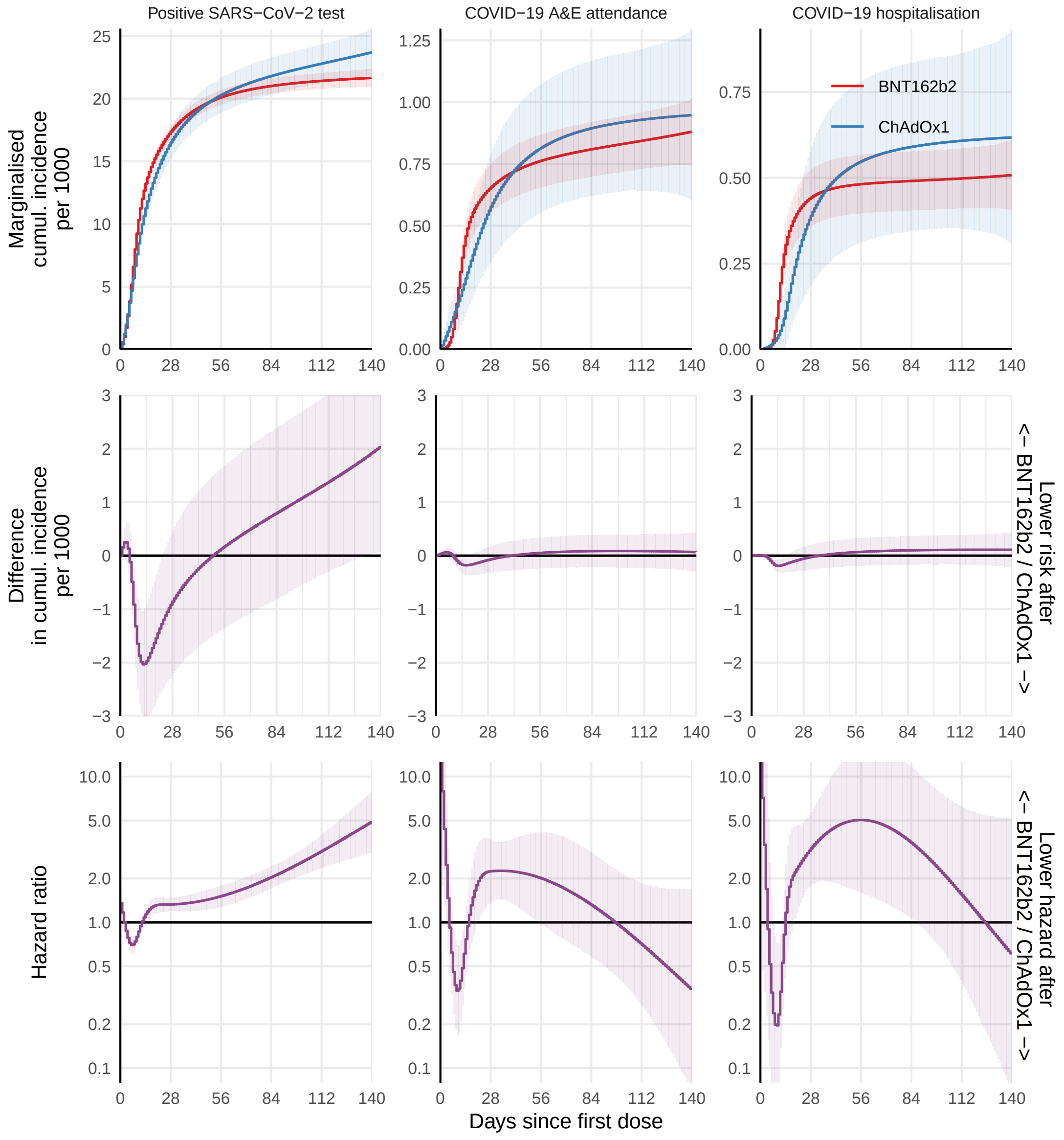

**Figure S2: Fully adjusted comparative effectiveness using piecewise-constant estimates of the hazard**

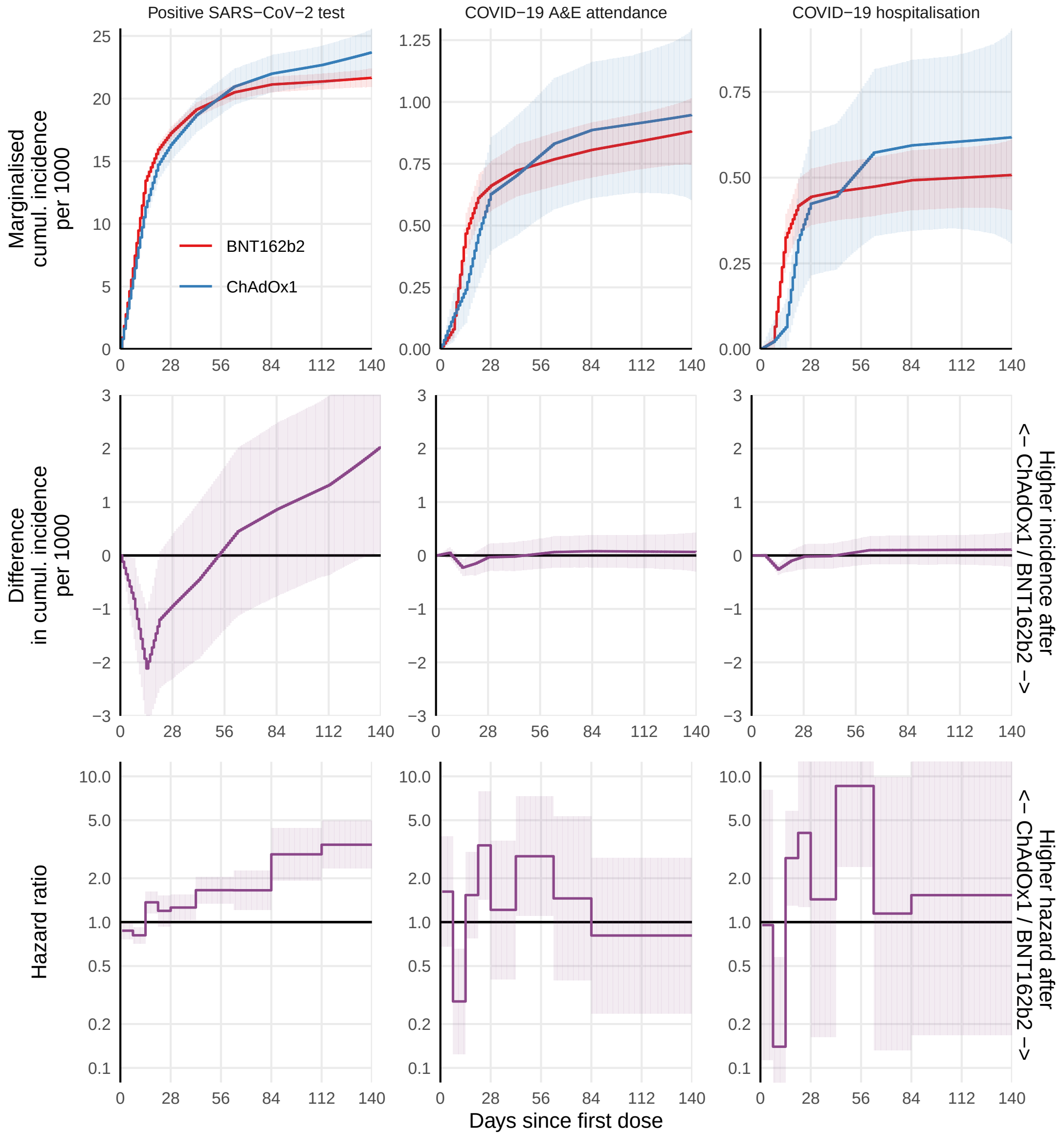

**Figure S3: Second dose interval**

Second dose events are censored by study end date, de-registration, death, and additionally by outcome-specific events.

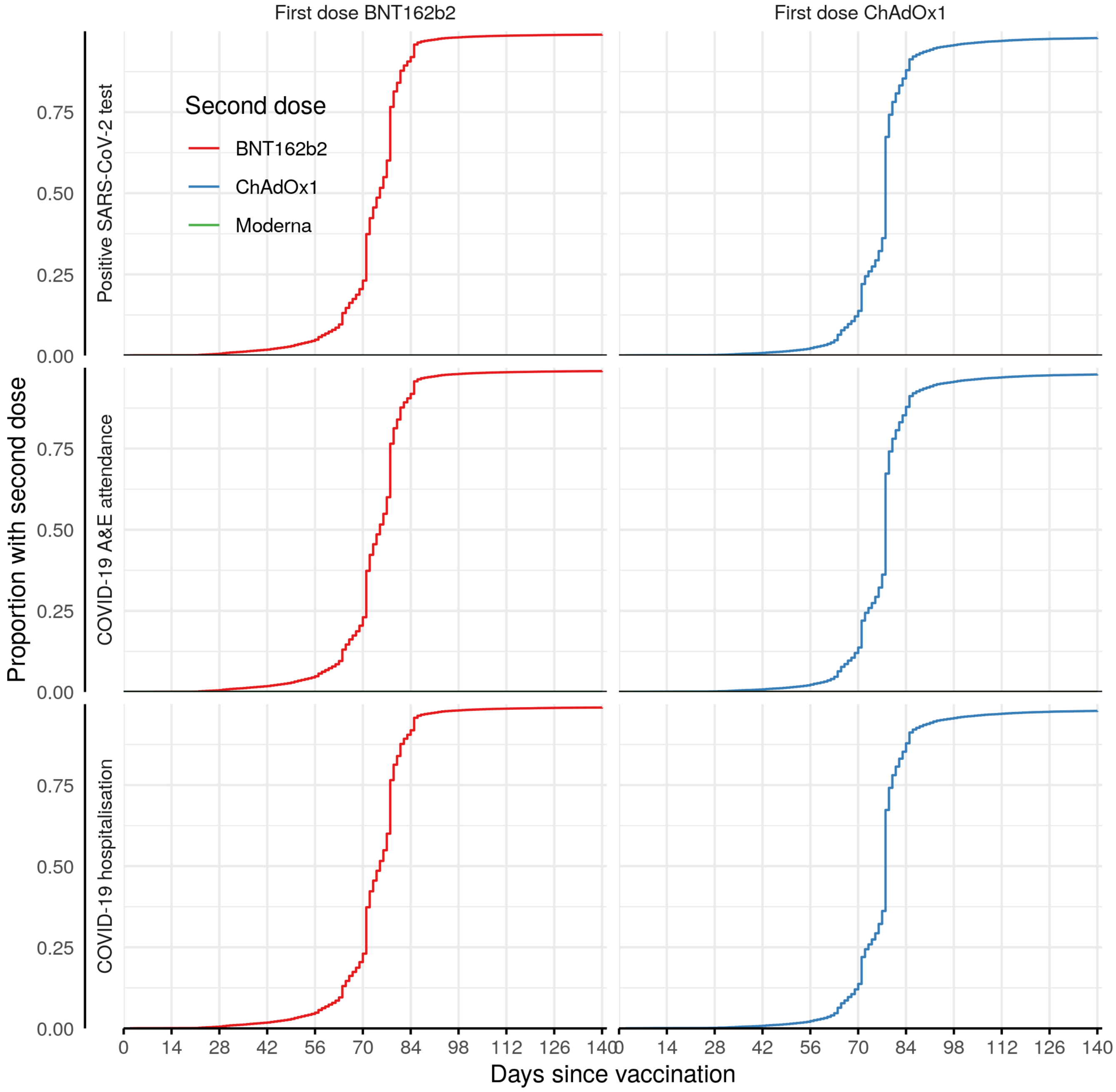

**Figure S4: Event rates by calendar time**

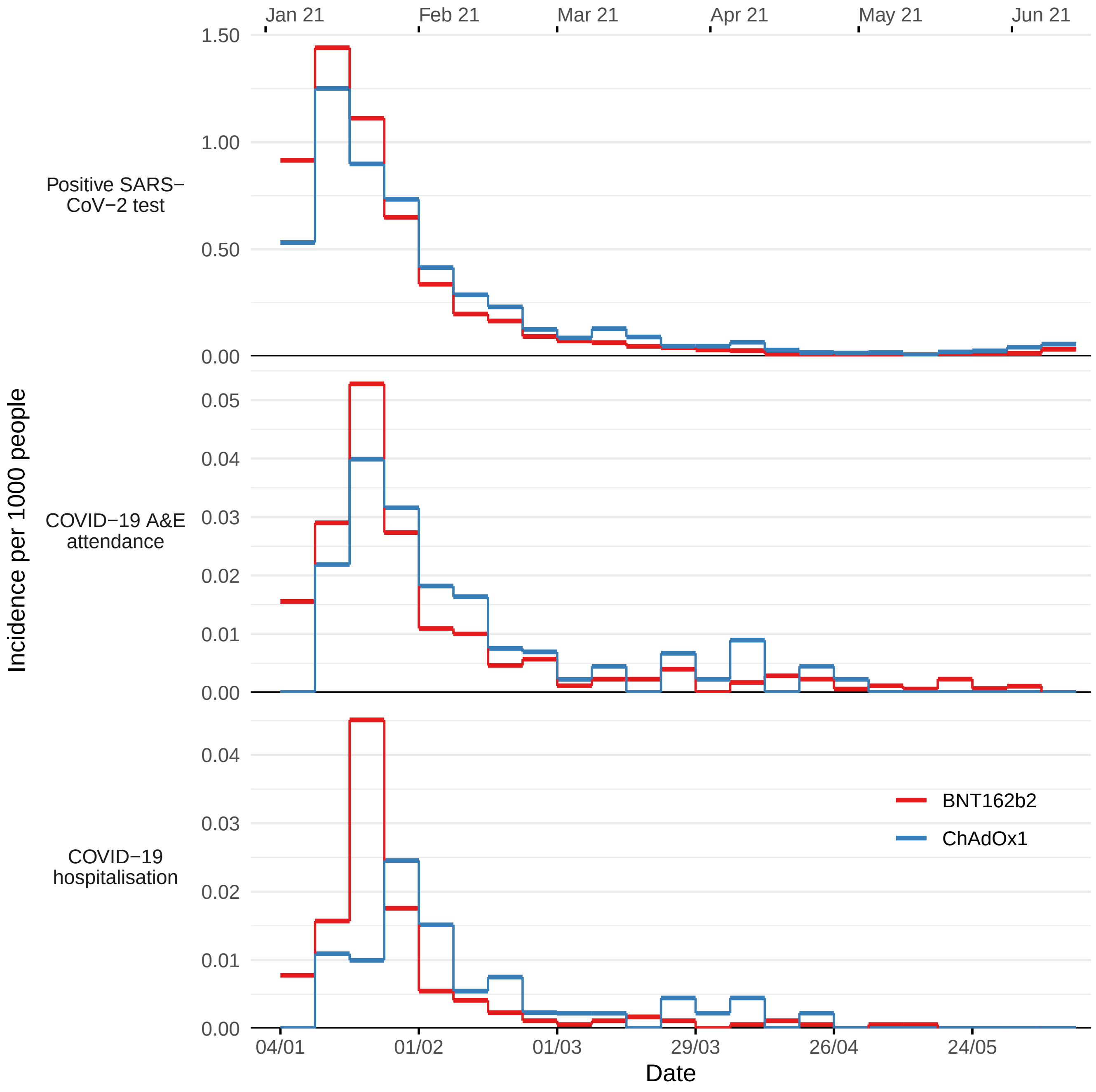

**Table S1: Inclusion criteria**

| Participants | N | N excluded | % remaining |
|---|---|---|---|
| HCWs aged 18-64 receiving BNT162b2 or ChAdOx1 between 4 Jan and 28 Feb | 361,287 | - | 100.0% |
| with no missing demographic information | 327,254 | 34,033 | 90.6% |
| who are not clinically extremely vulnerable | 317,341 | 9,913 | 87.8% |

**Table S2: Baseline characteristics before eligibility exclusions**

Characteristics for all HCWs aged 18-64 receiving a first dose of BNT162b2 or ChAdOx1 between 4 January and 28 February 2021 and actively registered at at TPP practice are reported, before exclusions due to missing demographic characteristics and Clinically Extremely Vulnerable status.

| Characteristic | BNT162b2 | ChAdOx1 |
|---|---|---|
| Total N | 286,559 | 74,728 |
| Age | | |
| 18-30 | 47,700 (17%) | 12,462 (17%) |
| 30s | 66,352 (23%) | 16,769 (22%) |
| 40s | 72,266 (25%) | 18,777 (25%) |
| 50s | 77,047 (27%) | 20,481 (27%) |
| 60-64 | 23,194 (8.1%) | 6,239 (8.3%) |
| Sex | | |
| Female | 225,335 (79%) | 57,290 (77%) |
| Male | [REDACTED] | [REDACTED] |
| Unknown | [REDACTED] | [REDACTED] |
| Ethnicity | | |
| White | 223,555 (83%) | 58,810 (84%) |
| Black | 9,381 (3.5%) | 3,707 (5.3%) |
| South Asian | 24,549 (9.2%) | 5,188 (7.4%) |
| Mixed | 4,123 (1.5%) | 1,145 (1.6%) |
| Other | 6,532 (2.4%) | 1,147 (1.6%) |
| Unknown | 18,419 | 4,731 |
| IMD | | |
| 1 most deprived | 40,175 (14%) | 11,653 (16%) |
| 2 | 51,761 (19%) | 13,652 (19%) |
| 3 | 61,325 (22%) | 15,420 (21%) |
| 4 | 63,102 (23%) | 16,116 (22%) |
| 5 least deprived | 61,237 (22%) | 15,362 (21%) |
| Unknown | 8,959 | 2,525 |
| Region | | |
| North East and Yorkshire | 60,707 (21%) | 19,091 (26%) |
| East of England | 70,597 (25%) | 14,081 (19%) |
| Midlands | 56,664 (20%) | 19,752 (26%) |
| South West | 41,161 (14%) | 5,981 (8.0%) |
| London | 11,672 (4.1%) | 2,502 (3.3%) |
| North West | 26,570 (9.3%) | 9,030 (12%) |
| South East | 19,088 (6.7%) | 4,268 (5.7%) |
| Unknown | 100 | 23 |
| Rural/urban category | | |
| Urban conurbation | 67,119 (24%) | 21,439 (30%) |
| Urban city or town | 158,696 (57%) | 36,582 (51%) |
| Rural town or village | 51,879 (19%) | 14,204 (20%) |
| Unknown | 8,865 | 2,503 |
| Vaccination day (from 4 January 2021) | 12 (7, 19) | 20 (13, 34) |
| Body Mass Index > 40 kg/m^2 | 11,102 (3.9%) | 3,421 (4.6%) |
| Chronic heart disease | 10,485 (3.7%) | 3,055 (4.1%) |
| Chronic kidney disease | 2,517 (0.9%) | 806 (1.1%) |
| Diabetes | 14,798 (5.2%) | 4,490 (6.0%) |
| Chronic liver disease | 4,426 (1.5%) | 1,416 (1.9%) |
| Chronic respiratory disease | 3,635 (1.3%) | 1,266 (1.7%) |

| | | |
|---|---|---|
| Chronic neurological disease | 6,863 (2.4%) | 1,941 (2.6%) |
| Immunosuppressed | 4,465 (1.6%) | 1,573 (2.1%) |
| Asplenia or poor spleen function | 2,169 (0.8%) | 654 (0.9%) |
| Learning disabilities | 215 (<0.1%) | 87 (0.1%) |
| Serious mental illness | 1,397 (0.5%) | 501 (0.7%) |
| Morbidity count | | |
| 0 | 235,874 (82%) | 59,642 (80%) |
| 1 | 42,344 (15%) | 12,174 (16%) |
| 2+ | 8,341 (2.9%) | 2,912 (3.9%) |
| Prior SARS-CoV-2 infection | 30,509 (11%) | 10,486 (14%) |
| Number of SARS-CoV-2 tests in previous 3 months | | |
| 0 | 182,942 (64%) | 49,148 (66%) |
| 1-3 | 82,447 (29%) | 21,857 (29%) |
| 4-6 | 11,988 (4.2%) | 2,099 (2.8%) |
| 7+ | 9,182 (3.2%) | 1,624 (2.2%) |
| Clinically Extremely Vulnerable | 6,838 (2.4%) | 3,578 (4.8%) |

**Table S3: Event rates**

| | BNT162b2 | | ChAdOx1 | |
|---|---|---|---|---|
| Days since first dose | Events / person-years | Incidence | Events / person-years | Incidence |
| Positive SARS-CoV-2 test | | | | |
| 1-14 | 21 / 103 | 0.204 | 16 / 72 | 0.221 |
| 15-28 | 26 / 102 | 0.255 | 24 / 72 | 0.335 |
| 29-42 | 14 / 101 | 0.138 | 8 / 71 | 0.113 |
| 43-70 | 54 / 199 | 0.271 | 42 / 139 | 0.302 |
| 71-84 | 29 / 98 | 0.296 | 14 / 68 | 0.205 |
| 85-98 | 25 / 97 | 0.258 | 20 / 67 | 0.296 |
| 99-112 | 24 / 94 | 0.255 | 17 / 66 | 0.259 |
| 113-126 | 17 / 77 | 0.220 | 15 / 56 | 0.269 |
| 127-140 | 14 / 54 | 0.258 | 14 / 41 | 0.345 |
| All | 224 / 926 | 0.242 | 170 / 652 | 0.261 |
| COVID-19 A&E attendance | | | | |
| 1-14 | – / 103 | – | – / 73 | – |
| 15-28 | 0 / 103 | 0 | – / 72 | – |
| 29-42 | – / 103 | – | 0 / 72 | 0 |
| 43-70 | – / 206 | – | – / 144 | – |
| 71-84 | – / 103 | – | 0 / 72 | 0 |
| 85-98 | 0 / 102 | 0 | 0 / 72 | 0 |
| 99-112 | 0 / 100 | 0 | – / 70 | – |
| 113-126 | 0 / 83 | 0 | 0 / 60 | 0 |
| 127-140 | – / 59 | – | 0 / 44 | 0 |
| All | 7 / 964 | 0.007 | 8 / 679 | 0.012 |
| COVID-19 hospitalisation | | | | |
| 1-14 | 7 / 103 | 0.068 | – / 73 | – |
| 15-28 | – / 103 | – | 6 / 72 | 0.083 |
| 29-42 | 6 / 103 | 0.058 | – / 72 | – |
| 43-70 | 7 / 204 | 0.034 | 12 / 143 | 0.084 |
| 71-84 | 6 / 102 | 0.059 | 6 / 71 | 0.085 |
| 85-98 | – / 102 | – | – / 71 | – |
| 99-112 | 9 / 99 | 0.091 | – / 69 | – |
| 113-126 | – / 82 | – | – / 59 | – |
| 127-140 | 6 / 58 | 0.103 | – / 43 | – |
| All | 54 / 956 | 0.056 | 42 / 673 | 0.062 |
| COVID-19 death | | | | |
| 1-14 | – / 103 | – | – / 73 | – |
| 15-28 | – / 103 | – | 0 / 73 | 0 |
| 29-42 | 0 / 103 | 0 | 0 / 72 | 0 |
| 43-70 | 0 / 206 | 0 | – / 144 | – |
| 71-84 | 0 / 103 | 0 | 0 / 72 | 0 |
| 85-98 | 0 / 103 | 0 | 0 / 72 | 0 |
| 99-112 | 0 / 101 | 0 | 0 / 71 | 0 |
| 113-126 | 0 / 84 | 0 | 0 / 60 | 0 |
| 127-140 | 0 / 59 | 0 | – / 44 | – |
| All | – / 965 | – | – / 681 | – |
| Second dose | | | | |

| | | | | |
|---|---|---|---|---|
| 1-14 | 0 / 103 | 0 | 0 / 73 | 0 |
| 15-28 | 0 / 103 | 0 | 0 / 73 | 0 |
| 29-42 | 0 / 103 | 0 | 0 / 72 | 0 |
| 43-70 | 0 / 206 | 0 | 0 / 144 | 0 |
| 71-84 | 109 / 102 | 1.066 | 70 / 72 | 0.978 |
| 85-98 | 283 / 93 | 3.029 | 217 / 65 | 3.323 |
| 99-112 | 293 / 80 | 3.642 | 178 / 57 | 3.146 |
| 113-126 | 127 / 59 | 2.145 | 78 / 43 | 1.796 |
| 127-140 | 0 / 41 | 0 | 0 / 31 | 0 |
| All | 812 / 893 | 0.910 | 543 / 630 | 0.862 |

**Table S4: Vaccine type for second doses**

| First dose | Matching second dose | N | % |
|---|---|---|---|
| BNT162b2 | no | 288 | 0.1% |
| BNT162b2 | yes | 249,091 | 98.4% |
| BNT162b2 | no second dose at 20 weeks | 3,755 | 1.5% |
| ChAdOx1 | no | 125 | 0.2% |
| ChAdOx1 | yes | 62,463 | 97.3% |
| ChAdOx1 | no second dose at 20 weeks | 1,619 | 2.5% |
